# Interpretable machine learning applied to high-dimensional salivary proteomics accurately classifies pediatric inflammatory bowel diseases

**DOI:** 10.1101/2025.10.14.25337919

**Authors:** Brittany T. Rupp, Joaquin Reyna, Ally Giunta, Theresa Weaver, Kelly Chason, Jinze Liu, Ajay S. Gulati, Kevin M. Byrd

**Author notes:** Authors with equal contribution (Brittany T. Rupp, Joaquin Reyna). CORRESPONDING AUTHORS: **Kevin Matthew Byrd, DDS Ph.D: Virginia Commonwealth University, VCU School of Dentistry, Philips institute of Oral Health Research**, Perkinson Building, Room 4130, 1101 East Leigh Street, P.O. Box 980566, Richmond, Virginia 23298-0566, USA, **E:**; **Ajay Sujan Gulati, MD: University of North Carolina, UNC School of Medicine**, 230 MacNider Building, CB# 7229, Chapel Hill, NC 27599-7593, USA, **E:**.

## Abstract

**Background and aims:** Inflammatory bowel diseases (IBD), including Crohn’s disease (CD), ulcerative colitis (UC), and IBD-unclassified (IBD-U), are chronic inflammatory disorders of the gastrointestinal tract. Current methods for classification and longitudinal monitoring are invasive, expensive, and often delayed, limiting timely diagnosis and management. This study reports the first application of high-dimensional salivary proteomics integrated with interpretable artificial intelligence/machine learning (AI/ML) to define a minimal protein signature for pediatric IBD classification with the goal of informing therapeutic decision-making.

**Methods:** Unstimulated saliva from pediatric CD, UC, and IBD-U patients was analyzed using Alamar Biosciences’ NULISAseq Inflammation Panel 250 (250 proteins). Logistic regression with recursive feature elimination identified a minimal discriminative signature. Performance was tested in independent follow-up samples. SHapley Additive exPlanations (SHAP) quantified patient-specific protein contributions and assessed biological similarity of IBD-U to CD and UC.

**Results:** Differential abundance analysis between UC and CD revealed 53 significantly different proteins. ML identified a 14-protein signature comprising chemokines/cytokines (CCL1, IFNA1;IFNA13, IL12p70, IL34, TNFSF11/RANKL), receptors/ligands (CD40LG, ICOSLG, IL1R2, IL17RA), structural/tissue-remodeling proteins (CD93, GFAP, SPP1), and growth factors/immune modulators (GDF2, GZMA). The model achieved 96.2% overall accuracy in first-visit samples and 86.4% overall accuracy in follow-up testing. SHAP revealed patient-specific drivers and suggested biological alignment of IBD-U cases toward CD-like or UC-like profiles.

**Conclusions:** This first-in-field integration of salivary proteomics with interpretable AI/ML demonstrates that accurate, noninvasive classification of pediatric IBD is possible using minimal biomarker sets. This approach establishes a scalable framework for future longitudinal monitoring, and supports earlier and more precise therapeutic interventions.

## INTRODUCTION

Inflammatory bowel diseases (IBD), encompassing ulcerative colitis (UC), Crohn’s disease (CD), and IBD-unclassified (IBD-U), comprise a group of chronic, relapsing conditions characterized by persistent, dysregulated immune responses and inflammation within the gastrointestinal tract^1,2^. In pediatric populations, early and accurate subtype classification is particularly critical, as ongoing inflammation during key developmental years can result in irreversible bowel damage, growth failure, delayed puberty, and reduced quality of life^3^. Despite advances in understanding IBD pathophysiology, current gold standard strategies for diagnosis and monitoring such as endoscopy and biopsy are invasive, costly, burdensome, and impractical for frequent longitudinal monitoring in children^4,5^. These limitations underscore the urgent need for precise, non-invasive biomarkers that can both distinguish CD from UC at diagnosis and support ongoing monitoring of disease activity and treatment response.

Biomarkers from human biofluids, including blood, plasma, urine, stool, sputum, and saliva, offer an attractive alternative approach for longitudinal monitoring of disease activity and drug response by enabling repeatable sampling in routine clinical settings. Each biofluid presents unique strengths and limitations depending on the disease context, clinical question, and patient population^6,7^. Saliva stands out for its rapid, minimally invasive collection with high patient acceptability and independence from specialized infrastructure or trained phlebotomists. These attributes make it particularly well-suited for pediatric applications^8^. Saliva harbors a diverse repertoire of biological analytes, including cells, nucleic acids, extracellular vesicles, metabolites, and proteins^9^. This molecular diversity positions saliva as a practical and informative window into systemic disease states, while its ease of collection enables longitudinal monitoring with minimal patient burden, supporting real-time assessment of disease activity, progression, and therapeutic response in chronic conditions. Furthermore, extraintestinal manifestations of oral inflammatory lesions are well documented in pediatric IBD cases^10^, and the establishment of an oral-systemic (i.e., gum-gut) link is increasingly being established mechanistically^11^.

Proteomics is emerging as a powerful strategy for biomarker discovery, combining the accessibility of biofluids with the ability to profile systemic inflammatory and immune pathways^12,13^. Recent advances, including high-sensitivity proximity ligation–based assays, enable multiplexed quantification of hundreds to thousands of proteins from extremely small sample volumes^14^. These advances permit comprehensive profiling of inflammatory networks from just a few microliters of biofluids like saliva. In parallel, the rapid emergence of artificial intelligence and machine learning (AI/ML) in proteomics is revolutionizing how complex, high-dimensional datasets are analyzed, facilitating the discovery of minimal yet highly discriminative biomarker panels, improving disease classification accuracy and enhancing interpretability to support clinical decision-making^15^. Adult IBD studies have applied such platforms to explore a variety of factors including need for treatment escalation^16^, chance of developing IBD^17^, ability to distinguish CD from UC (AUC=0.73)^18^, and disease location^19^.

These adult IBD models have limited applicability to pediatric patients due to differences in disease etiology, immune maturation, phenotypic presentation, and the higher prevalence of IBD-unclassified (IBD-U) in children^20,21^. To address this gap, we piloted salivary proteomics using the NULISAseq platform for the first time in a clinical research setting through the IBD-SCAN study at the University of North Carolina’s Multidisciplinary Pediatric IBD Center. To maximize downstream utility, we implemented a multi-tube collection protocol, allowing each sample to be split and stored for proteomics, microbiome profiling, cell-based immune and epithelial assays, and other -omics applications. This design not only addressed the study’s primary aims but also established a versatile salivary biorepository to support future investigations into the molecular underpinnings of pediatric IBD.

In this study, one aliquot of each saliva sample was analyzed using the NULISAseq Inflammation Panel 250, which provides high multiplexing capacity and exceptional sensitivity for low-abundance targets. This enabled the quantification of 250 inflammation-associated proteins directly from saliva. We then applied an interpretable machine learning pipeline to identify a minimal yet highly discriminative protein signature capable of differentiating CD from UC. Model interpretability was enhanced through SHapley Additive exPlanations (SHAP), which quantified the contribution of each protein to classification and enabled biological comparisons between IBD-U cases and CD/UC proteomic profiles. By combining next-generation salivary proteomics with state-of-the-art AI/ML tools, our approach demonstrates the potential for precision, non-invasive disease classification that is not only accurate but also clinically interpretable, which are key factors for eventual integration into pediatric IBD care.

## Materials and Methods

### Participants

Saliva samples were collected from IBD patients at the University of North Carolina (UNC) Hospitals Children’s Specialty Clinic. This study aimed to recruit both male and female patients ages seven to twenty-one years old from all races and ethnicities. Patients and/or legal guardians provided informed consent for sample collection, which was part of a protocol that obtained ethical approval from the UNC Institutional Review Board (IRB) (UNC #23-2319; Inflammatory Bowel Disease Salivary Cell Analysis Network; IBD-SCAN). All patients were 21 years old or younger at the time of collection and were 18 years old or younger at the time of IBD diagnosis. Patient’s clinical and demographic data is provided in table 1. Samples were collected from patients with Crohn’s disease (CD), ulcerative colitis (UC), or IBD-U (IBD Unclassified). Follow-up samples were collected from the same IBD patient if the patient returned to the clinic during the collection period (7.5 months).

**Table 1:** Patient Clinical and Demographic Data. Comparison of Crohn’s and ulcerative colitis patient cohort’s demographic and clinical characteristics. For the p-value column, ^ indicated Fisher Exact test was used to assess for statistically significant differences between cohorts. t indicates Mann-Whitney test was used.

| Characteristics | Crohn's Disease<br>(CD) | Ulcerative Colitis<br>(UC) | P value |
| --- | --- | --- | --- |
| Demographic |  |  |  |
| Total no. of patients, n (%) | 40 (76.9) | 12 (23.1) | - |
| Female, n (%) | 15 (37.5) | 5 (41.7) | 1.0000<sup>^</sup> |
| Age at first donation, y, mean (SD) | 16.1 (3.4) | 15.9 (2.7) | 0.6574<sup>†</sup> |
| Race, n (%) |  |  |  |
| American Indian/ Alaska Native | 0 (0.0) | 1 (8.3) | 0.2308<sup>^</sup> |
| Black/African American | 6 (15.0) | 2 (16.7) | 1.0000<sup>^</sup> |
| Caucasian | 25 (62.5) | 7 (58.3) | 1.0000<sup>^</sup> |
| Multiracial, other or unknown | 9 (22.5) | 2 (16.7) | 1.0000<sup>^</sup> |
| Ethnicity, n (%) |  |  |  |
| Hispanic or Latino | 1 (2.5) | 1 (8.3) | 0.4118<sup>^</sup> |
| Not Hispanic or Latino | 36 (90.0) | 11 (91.7) | 1.0000<sup>^</sup> |
| Unknown | 3 (7.5) | 0 (0.0) | 1.0000<sup>^</sup> |
| Clinical |  |  |  |
| Height, z score, mean (SD) | -0.029 (0.948) | 0.036 (0.829) | 0.5970<sup>†</sup> |
| Weight, z score, mean (SD) | 0.288 (1.010) | 0.151 (1.002) | 0.5903<sup>†</sup> |
| Body mass index, z score, mean (SD) | 0.354 (1.200) | 0.178 (0.891) | 0.4511<sup>†</sup> |
| Time from diagnosis, mo, mean (SD) | 51.8 (38.8) | 57.3 (27.6) | 0.4102<sup>†</sup> |
| Visit number, n (%) |  |  |  |
| 1 | 40 (71.4) | 12 (66.7) | 0.7699<sup>^</sup> |
| 2 | 11 (19.6) | 3 (16.7) | 1.0000<sup>^</sup> |
| 3 | 5 (8.9) | 2 (11.1) | 1.0000<sup>^</sup> |
| 4 | 0 (0.0) | 1 (5.6) | 0.2432<sup>^</sup> |
| Physician Global Assessment, n (%) |  |  |  |
| Quiescent | 32 (80.0) | 9 (75.0) | 0.7012<sup>^</sup> |
| Mild | 7 (17.5) | 1 (8.3) | 0.6634<sup>^</sup> |
| Moderate | 1 (2.5) | 2 (16.7) | 0.1294<sup>^</sup> |
| Latest IBD Treatment, n (%) |  |  |  |
| Anti TNF | 26 (63.4) | 1 (8.3) | 0.0007<sup>^</sup> |
| Corticosteroids | 6 (14.6) | 1 (8.3) | 1.0000<sup>^</sup> |
| Immunomodulators | 2 (4.9) | 1 (8.3) | 0.5529<sup>^</sup> |
| Anti-IL-23 | 1 (2.4) | 1 (8.3) | 0.4118<sup>^</sup> |
| Anti-Integrin | 2 (4.9) | 3 (25.0) | 0.0739<sup>^</sup> |
| JAK Inhibitor | 1 (2.4) | 2 (16.7) | 0.1294<sup>^</sup> |
| 5-Aminosalicylate | 0 (0) | 1 (8.3) | 0.2308<sup>^</sup> |

### Saliva collection and processing

Before collection, patients were advised not to consume foods with a high acidity, caffeine, or sugar content, and to avoid eating a major meal within 60 minutes of sample collection. One or more milliliters of unstimulated saliva was collected from each patient via passive drool. In brief, patients were instructed to sit for 3-5 minutes without swallowing to allow saliva to pool in their mouth. After saliva was pooled, the patient would open their mouth and push the saliva into an open conical tube. Once collected, samples were placed on ice until they could be processed and frozen. For processing, the conical tubes containing the samples were centrifuged at 300g for 5 minutes. The supernatant was removed without disturbing the cell pellet and placed in a labelled cryovial. Cryovials were cataloged and frozen at -80C for long term storage.

### Measurement of Inflammatory Proteins in Samples

Saliva supernatant samples were shipped overnight on dry ice to Alamar Biosciences to analyze for 250 inflammation related proteins using the NULISAseq Inflammation Panel 250 (supplementary table 1). In brief, 30ul from 82 samples was placed in a 96 well plate along with sample controls, inter-plate controls and negative controls provided by Alamar. Specific, individual proteins were detected using dual antibody selection proximity ligation which generated a DNA reporter molecule containing a unique pair of target-specific molecular identifiers (TMIs). Sample-specific molecular identifiers were also introduced to the DNA reporters to allow for multiple samples to be pooled for next generation sequencing. After sequencing, raw sequencing reads were processed to generate target specific counts for each sample. These counts were normalized according to Alamar’s protocol using the internal control and inter-plate controls run simultaneously with the IBD samples. Samples counts were divided by the internal control counts and the inter-plate control medians before being rescaled and transformed into NULISA Protein Quantification (NPQ) units. NPQ units were used for the majority of analysis. When fold change is reported, it is calculated as 2^(Difference in NPQ)

### Sample visualization using principal component analysis and heatmaps

To explore the dataset, principal component analysis (PCA) was performed using all CD and UC samples with different subsets of proteins (e.g., all 250 proteins, 53 differentially abundant proteins). To visualize the principal component space for each disease subgroup, a scatterplot was drawn with convex hulls plotted around the corresponding points (using scipy.spatial.ConvexHull)^22^. In addition, heatmaps with hierarchical clustering were produced with the ComplexHeatmaps package in R^23^. Prior to plotting, the levels of each protein were mean-centered and scaled to unit variance using the scale() function in R, and the resulting values were used as input.

### Differential Abundance Analysis

Samples were filtered to include only participants diagnosed with either CD or UC, using all 250 proteins as features. Differential abundance analysis was performed using the limma package in R (version 4.3.3)^24^. As NPQ values were derived from log2-transformed values, they were directly used as input. To account for repeated measures, the duplicateCorrelation() (from limma) was used to calculate intra-patient correlation and these values were incorporated during the linear modeling steps of limma. From the limma output, log2 fold changes (logFC) and raw p-values were extracted. Multiple testing correction was applied using the Benjamini-Hochberg procedure. Although FDR-adjusted P-values were computed, few proteins met the conventional significance threshold (FDR < 0.05), likely due to small sample size and inherent variability of proteomics data. Therefore, for exploratory purposes, proteins with nominal P-values < 0.05 were reported and used for downstream analysis, with appropriate caution regarding potential false positives. Volcano plots were also generated, with reported proteins colored by directionality and the top proteins (ranked by nominal P-value) labeled.

### Model Development Using Recursive Feature Elimination and Application

For model development, the 82 samples were divided into three subgroups:

- Training data: Samples derived from each patient’s first visit and diagnosed with CD (n=40) or UC (n=12) only, used exclusively and strictly for model training and validation.
- Testing data: Samples from subsequent visits (post visit 1) and diagnosed with CD (n=16) or UC (n=6) only, used exclusively for final model testing.
- IBD-U only: Samples from patients with an undiagnosed IBD subtype (n=8), used for novel inference only.

#### i. Prototyping Logistic Regression Models

A prototyping dataset was generated from the training data using an 80/20 split and all 250 proteins as features. To correct for class imbalance, UC samples were randomly oversampled until the number of UC observations matched those of CD^25^. Logistic regression models were trained using default parameter settings (including a preset L2 norm) as implemented in sklearn’s LogisticRegression() function ^26^. To survey a variety of logistic regression models, we employed recursive feature selection using sklearn’s RFE() function while varying the number of selected features^26^. Model performance was evaluated using F1 scores.

#### ii. Validation of Models with Leave-One-Out Cross-Validation

For each RFE model, leave-one-out validation was performed, and performance metrics were collected at each fold. For ROC curve analysis, class probabilities P (CD=1 | features) were estimated from the logistic regression to calculate the false positive rate, true positive rate, and area under the curve (AUC). Confusion matrices for the top-performing models were generated using the same leave-one-out approach by comparing predicted and true labels to derived counts for each quadrant. Accuracy, precision, recall, and F1 scores were also calculated to further evaluate model performance.

#### iii. Model Testing Using Post-V1 and IBD-U Samples

To further assess the predictive performance of different RFE models, all samples corresponding to secondary visits (post visit 1) were extracted and predictions were made. As described previously, performance was assessed using calculations for accuracy, precision, recall, F1 scores, and confusion matrices and a final model was selected. The optimized RFE model was then utilized to generate predictions for IBD-U samples and further investigated in downstream analyses.

### Assessing Target Protein Detectability and Predictive Potential

To test protein detectability relative to the lower limit of detection (LOD), the protocol described by Alamar was utilized^14^. Briefly, the protein level of a given sample was compared to its corresponding lower limit of detection and categorized as either above or below this threshold. A target protein was then considered detectable if more than 50% of samples exceeded the LOD threshold. To evaluate the robustness of this protocol, model development using recursive feature elimination was repeated using detectable proteins (n=230) and the resulting RFE logistic regression models were compared using the confusion matrix, accuracy, precision recall and F1 scores, as described previously.

### Model Explanation Using SHAP Value Analysis

To interpret the final RFE logistic regression model and individual predictions, SHAP (SHapley Additive exPlanations) analysis was conducted using the SHAP Python package^27^. Since the logistic regression is a linear model, the linear explainer (shap.LinearExplainer) was employed for efficient and exact computation of SHAP values. Model explanation was performed in three phases:

- Training data (Visit 1 samples): SHAP values were computed for the visit 1 dataset, and global feature importance was analyzed by averaging SHAP contributions for each protein across all samples. To visualize these results, the shap.plots.bar function was used.
- Testing data (Post Visit 1 samples): SHAP values were similarly calculated for post visit 1 samples. Force plots were generated (shap.plots.force) for select samples to visualize how specific proteins contribute to high-confidence predictions (i.e. strong CD or UC classifications) as well as ambiguous predictions (log-odds close to zero).
- Inference on novel data (IBD-U samples): For the IBD-U subset, force plots were generated to visualize contributions in both strong and ambiguous predictions. A decision plot (shap.decision_plot) was also generated to provide a cohort-level overview of protein contributions and their cumulative influence on prediction outcomes across all samples.

### Graph generation and statistical analysis

All box plots, bar graphs and heatmaps without hierarchical clustering were generated using GraphPad Prism V10.4.1. For Figure 2 and supplemental figure 2, box plots contain values from all CD and UC samples, and reported p-values were generated as part of the Differential Abundance Analysis. For Figure 4b, graphs contained CD and UC values from V1 samples only, and p-values were generated using a Mann-Whitney test in GraphPad Prism.

## RESULTS

### IBD-SCAN Patient Cohort and Sample Collection

The persistent need for non-invasive, biomarkers for pediatric inflammatory bowel diseases (IBD) motivated the design of a prospective clinical study called IBD-SCAN, which was conducted under an Institutional Review Board–approved protocol at the University of North Carolina at Chapel Hill (IRB #23-2319). This study was conceived to address the challenge of accurately distinguishing Crohn’s disease (CD) from ulcerative colitis (UC) without relying solely on invasive endoscopic procedures. Saliva was selected as the biospecimen of choice due to its painless collection, minimal volume requirements, and capacity to capture both local and systemic inflammatory signals. The study design incorporated a multi-tube collection protocol to support parallel analyses across proteomics, microbiome profiling, and cell-based assays, thereby creating a versatile resource for both the primary investigation and future research. The central goal of IBD-SCAN was to determine whether advanced, integrated, and multiomic profiling combined with interpretable machine learning could generate robust, disease-specific signatures suitable for initial classification and longitudinal monitoring.

For this first cohort, a total of 52 pediatric IBD patients, aged 8–21 years at the time of collection and all diagnosed at or before the age of 18, were enrolled. The cohort included both CD and UC patients, with additional demographic and clinical details provided in Table 1. As indicated, there were no significant demographic differences between the two groups, with the only clinical distinction being a higher frequency of anti-TNF prescription in CD, consistent with standard therapeutic patterns. To capture temporal variation, patients returning for routine clinic visits during the study period contributed additional follow-up samples, with up to four timepoints per individual. In total, 74 saliva samples were collected from CD or UC patients, enabling both cross-sectional classification analyses and longitudinal assessments of proteomic stability and disease-associated shifts over time (i.e., visit 1, visit 2, etc.). A minimum of 250μl of saliva was collected from all patients (see: *Methods*).

### Pilot Evaluation of Salivary Proteomics using the NULISAseq Platform

We decided to use NULISAseq because it enables high-plex, ultra-sensitive protein detection from minimal sample volumes, making it well-suited for pediatric saliva samples where collection yield can be limited. Importantly, the platform’s 250-plex inflammation protein panel provided broad, quantitative coverage of relevant pathways, while its reproducibility and wide dynamic range allowed for robust detection of both low- and high-abundance proteins in a single assay, maximizing our ability to capture disease-relevant signals for IBD classification (Figure 1a, Supplementary Table 1). Before initiating large-scale processing and analysis in our pediatric IBD cohort, we first evaluated the feasibility and optimal parameters for applying the NULISAseq platform to saliva, an application that had not been previously reported for this assay (though it has been utilized for other comparable systems^14^). To accomplish this, saliva was collected from five healthy adult volunteers and processed to obtain cell-free supernatant through centrifugation. This fraction was used for proteomic analysis on Alamar Biosciences’ NULISAseq platform and was designed to determine the saliva input volume required to achieve robust protein detection while preserving the ease of collection that makes saliva particularly suitable for clinical and pediatric use.

**Figure 1:**
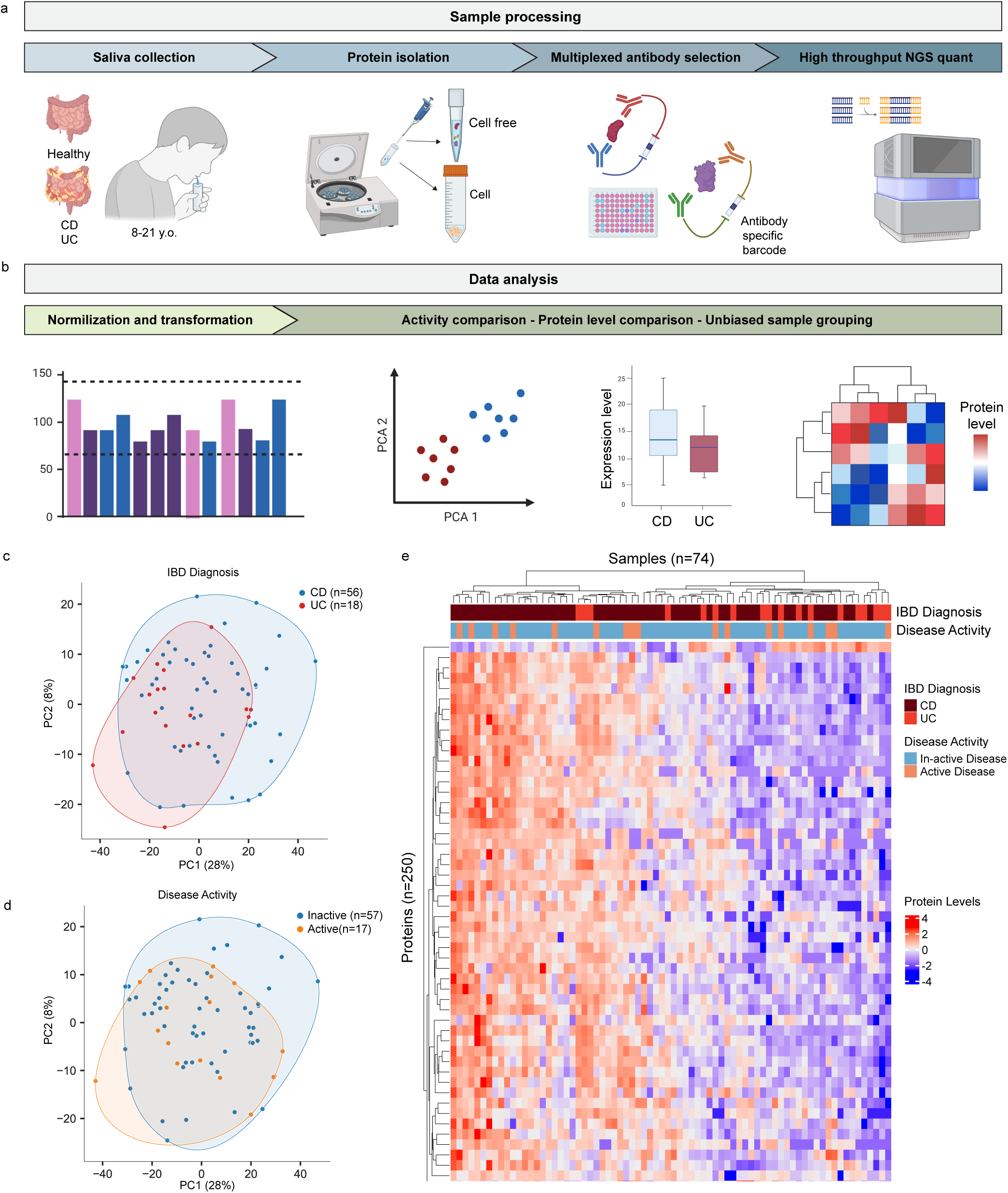
Study overview with full proteomic panel comparisons. a) Diagram depicting sample collection and processing workflow. b) Diagram depicting data analysis methods and pipeline. c) Principal component analysis of all Crohn’s (CD) and ulcerative colitis (UC) samples, colored by IBD diagnosis. d) Principal component analysis of all samples, colored by disease activity. e) Heatmap with hierarchical clustering of samples showing all 250 standardized protein levels from the NULISAseq Inflammation Panel 250. Corresponding IBD diagnosis and disease activity for each sample is shown. Illustrations from a) and b) were created with BioRender.com

We compared two input volumes, 10μl and 30μl, from the same donors to assess protein detection efficiency within a clinically practical range. On average, 197 proteins were detected at or above the limit of detection (LOD) in the 10μl samples, compared with 215.8 proteins in the 30μl samples, representing a 9.5% improvement in coverage with the larger volume (Supplementary Figure 1). Given that the increase in volume did not impose an additional collection burden, 30μl was selected for all subsequent analyses, including the pediatric IBD cohort. Only two proteins were undetectable in all healthy samples, and both were later detected in pediatric IBD saliva, suggesting that inflammatory disease states may increase the salivary abundance of certain proteins. This finding demonstrates that NULISA-based salivary proteomics can capture disease-associated changes in the inflammatory proteome, supporting its potential as a non-invasive tool for IBD classification and monitoring.

### Global Proteomic Patterns in Pediatric IBD

Salivary samples from the IBD-SCAN cohort were analyzed using the NULISAseq Inflammation Panel 250, with NULISA Protein Quantification (NPQ) units providing a relative measure of protein abundance across samples (see: *Methods,* Figure 1b). Principal component analysis (PCA) of the full 250-protein dataset revealed no clear separation by diagnosis (CD vs. UC) or by clinically assessed disease activity, and hierarchical clustering likewise showed only modest grouping by diagnosis (Figure 1c–e). These findings likely reflect the complexity of the salivary proteome, which is influenced by systemic inflammation as well as local oral factors. Interestingly, while fecal calprotectin may be a reliable biomarker for IBD activity, salivary S100A9 (component of calprotectin) did not correlate with disease activity in this cohort, suggesting the need to identify alternate biomarkers within the saliva IBD proteome (Supplemental Figure 2)^28^.

Including all 250 proteins in our analysis, some of which may be responsive to unrelated oral conditions, risked diluting disease-relevant signals. To focus on proteins most associated with IBD subtype differences, we identified 53 proteins with significantly different abundances between CD and UC (Figure 2a). Fifty-two were higher in CD, while only one (MPO) was enriched in UC (Figure 2b). Figure 2c shows box plots of the top ten differentially higher proteins in CD and the two differentially higher proteins in UC (the remaining 42 box plots can be found in Supplementary Figure 3). The range of protein levels varied across proteins, with some proteins (ex: CCL1, IFNA1;IFNA13) being undetectable in a portion of the samples. Restricting PCA and clustering to this subset improved separation by diagnosis, with the first principal component explaining 52% of the variance compared to 26% in the full dataset (Figures 2d–e). The CD-enriched proteins represented diverse immune and inflammatory pathways, including cell growth and differentiation factors (e.g., FGF2, GDF15, LIF), proinflammatory cytokines and chemokines (e.g., CCL1, CCL3, CXCL1, IL17A, IL22), type I/III interferons (e.g., IFNA1, IFNL1, IFNL2), immune cell costimulatory molecules (e.g., CD80, TNFRSF8), and mediators of matrix remodeling and barrier function (e.g., MMP3, LAMP3). Several of these pathways, including interferon signaling, IL-17–driven inflammation^29^ and TNF receptor family activation^30^, are established features of CD pathogenesis, while others, including the enrichment of growth factor signaling (FGF2, FGF23, LIF) in saliva, are less well characterized in pediatric IBD and may represent novel biological features. In UC, MPO may reflect selective enrichment of mucosal neutrophil. While this targeted assay is not an absolute quantification method, the consistent relative differences across subtypes suggest that salivary proteomics can sensitively capture disease-associated pathway activity in pediatric IBD.

**Figure 2:**
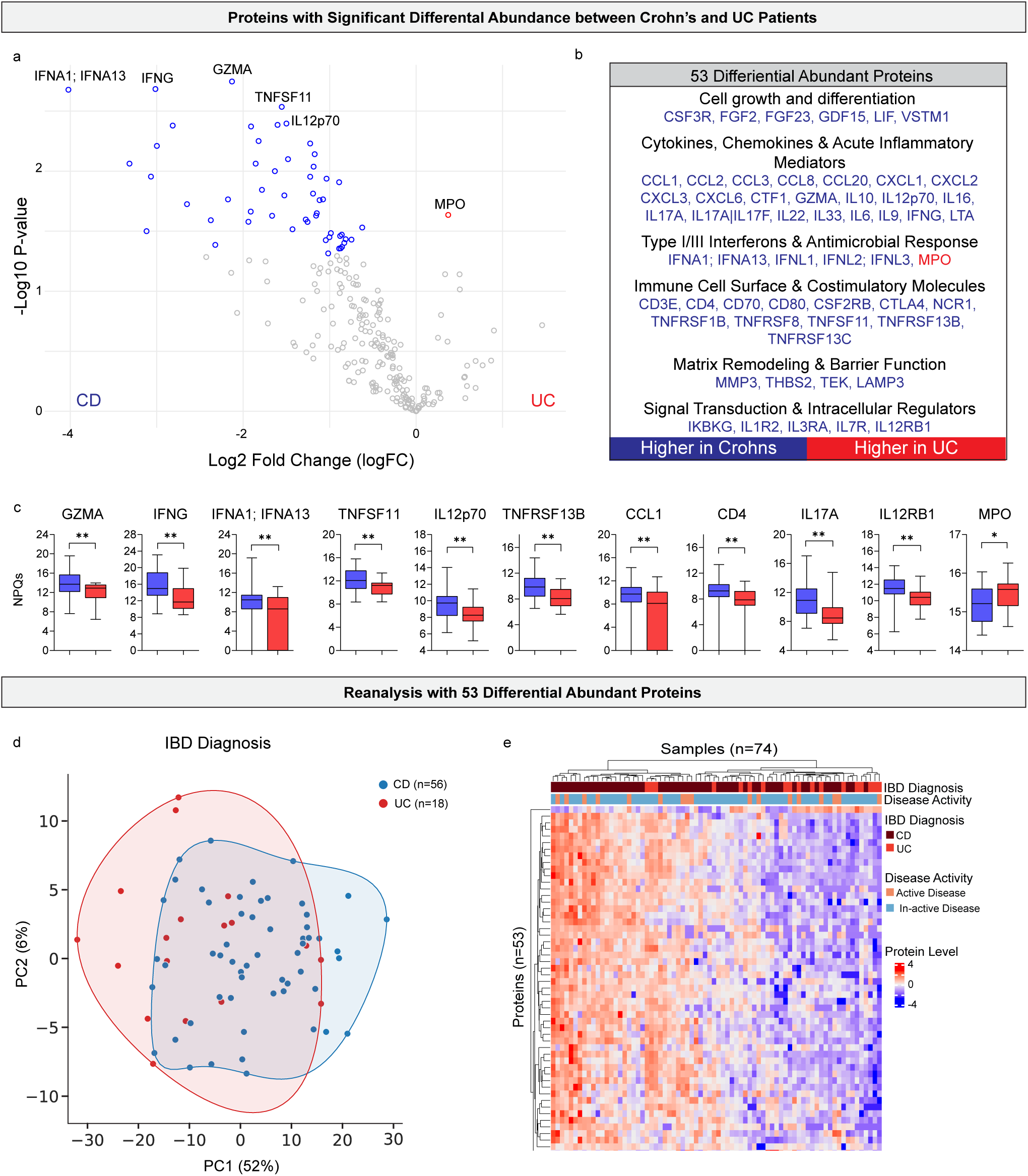
Comparison of proteomic signature differences between Crohn’s Disease and ulcerative colitis patients. a) Volcano plot comparing single proteomic markers between ulcerative colitis (UC), indicated by positive fold change (red), and Crohn’s disease (CD), indicated by negative fold change (blue). All colored dots indicate a p-value of <0.05. b) List of all statistically significant (p-value <0.05) differentially abundant proteins between UC and CD samples. c) Box plots of a selection of the most differentially abundant proteins by p value. Box plots show the distribution of protein levels (NULISA protein quantification units (NPQ)) in all CD patient samples (blue, n=56) and UC patient samples (red, n=18). Significance was determined using differential abundance analysis. * p value <0.05, ** p value <0.01. d) Principal component analysis of CD and UC samples using only statistically significant differentially abundant proteins listed in panel b, colored by IBD diagnosis. e) Heatmap with hierarchical clustering of CD and UC samples using only differentially abundant proteins listed in panel b. Protein levels were standardized, and corresponding IBD diagnosis and disease activity for each sample is shown in the upper bars.

### Machine Learning Classification of IBD Subtypes

Building on this, we then applied a logistic regression model with recursive feature elimination (RFE) to determine whether machine learning could better resolve subtype-specific patterns. To avoid bias toward individuals with multiple timepoints, only first-visit samples (n = 52) were included in the training set and initial model generation, with all remaining samples (n = 22) reserved for later testing. We applied RFE to identify a more informative protein signature by iteratively removing the least informative protein (those with the smallest model coefficients) and retraining the model at each step (Figure 3a). We initially prototyped models using an 80/20 split of first-visit samples, with evaluation metrics including F1 scores for both training and validation datasets (Figure 3b). These results demonstrated that salivary proteomics have strong predictive power, motivating us to further assess the model’s performance using leave-one-out cross-validation in all downstream analyses (Figure 3c-e). High performance was maintained across models containing 9–20 proteins, with F1 scores between 0.98 and 1.00, and Receiver Operating Characteristic (ROC) curve values between 0.91 and 0.99. Given the importance of minimizing signature size for clinical applicability, these results indicated that accurate classification could be achieved with fewer than 20 proteins. Confusion matrices comparing model-predicted versus clinician-assigned diagnoses (Figure 3d) revealed that the 15-protein model achieved the highest accuracy (98.1%), misclassifying only a single CD case as UC. This performance suggests that a small, well-curated protein panel can serve as the basis for a non-invasive, saliva-based diagnostic tool capable of distinguishing IBD subtypes with near-clinical accuracy (Figure 3e).

**Figure 3:**
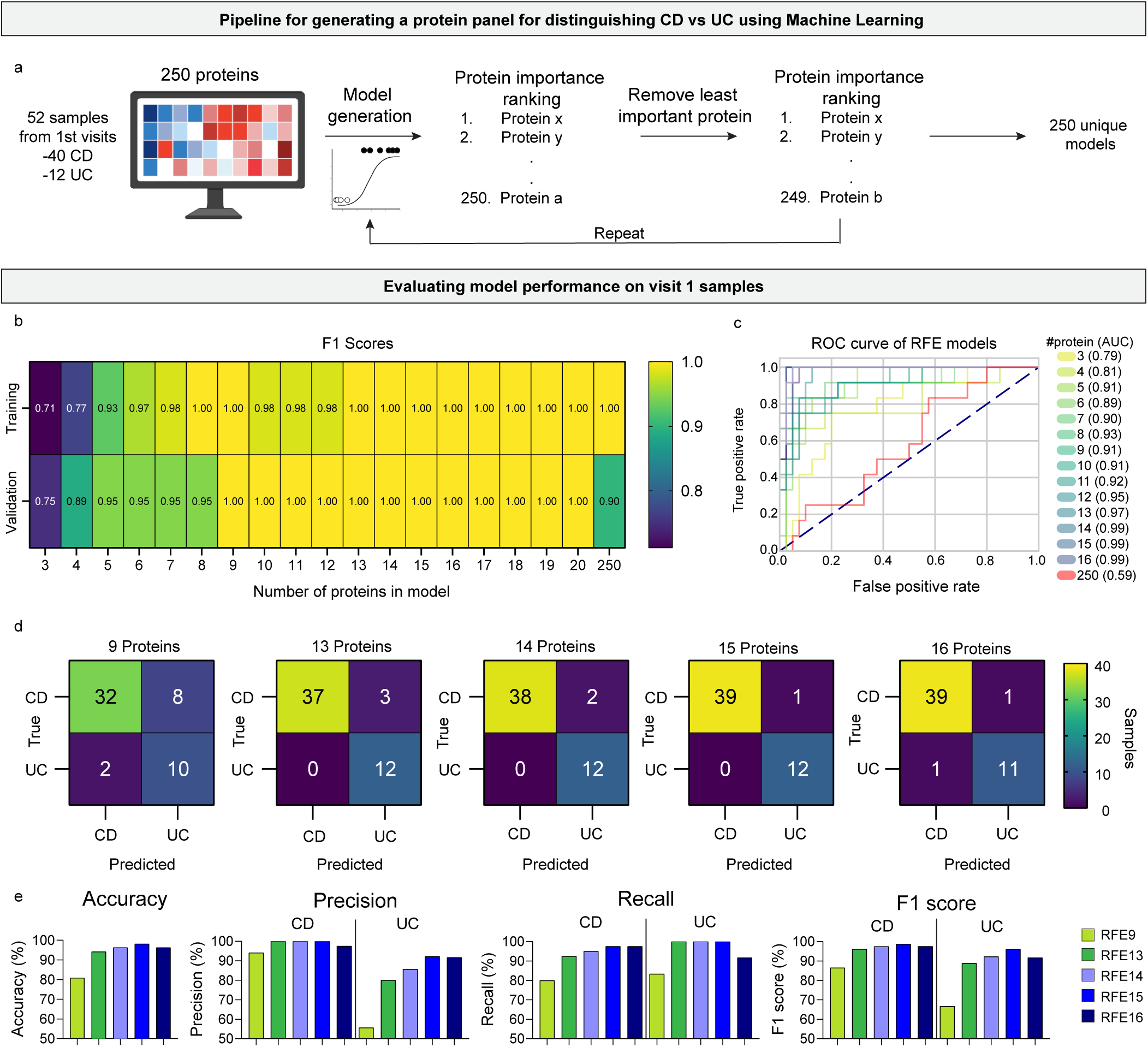
Development and evaluation of machine learning classification of CD and UC using salivary proteomics. a) Schematic overview of the recursive feature elimination (RFE) workflow used to develop a logistic regression model for classifying IBD subtypes. The model was trained on first-visit saliva samples (40 CD, 12 UC; total n = 52) to avoid bias from repeated measures. Proteins were ranked by importance, the least informative protein was removed, and the model was retrained at each step to generate a new unique model. b) Heatmap of the prototype models’ F1 score for training (top row) and validation (bottom row) datasets across protein panel sizes ranging from 3 to 250. c) Receiver operating characteristic (ROC) curves and area under the curve (AUC) values for selected RFE models using leave-one-out cross-validation. d) Confusion matrices for models containing 9, 13, 14, 15, and 16 proteins, with clinician IBD diagnosis (true) and model IBD diagnosis (predicted) indicated for visit 1 data. e) Summary bar charts of accuracy, precision, recall, and F1 scores for each model when applied to visit 1 data. Illustrations from a) were created with BioRender.com.

We additionally created a filtered quality-controlled dataset of 230 proteins, excluding proteins below the limit of detection in more than 50% of samples, and ran it in parallel to compare its performance with the full 250 protein model (Supplementary Figure 4). The QC-filtered model performed slightly less well (maximum accuracy 92.3%) than the unfiltered model (maximum accuracy 98.1%), demonstrating that our method does not require additional post-normalization QC. The unfiltered model was therefore selected for all downstream analyses to maximize discriminatory power.

### Testing and Interpretation of the Reduced Protein Model

While the initial machine learning results were strong, the model had been generated exclusively from first-visit data and therefore required further evaluation to assess its robustness on unseen samples. We tested its performance using follow-up visit data not included in the training process, finding that maximum overall accuracy remained high at 86.4% for the RFE13 and RFE14 with a slightly lower accuracy in RFE15 (81.8%, Figure 4a-b). Therefore, by comparing results from both initial and follow-up datasets, we determined that a 14-protein panel (RFE14) achieved the optimal balance between predictive accuracy and signature size, making it the most suitable model for downstream analyses. Notably, recall for ulcerative colitis remained modest at 66.7%, likely reflecting the lower representation of UC cases in the training set (Figure 4b). Increasing the number of UC samples in future cohorts should improve classification performance and reduce this imbalance.

**Figure 4:**
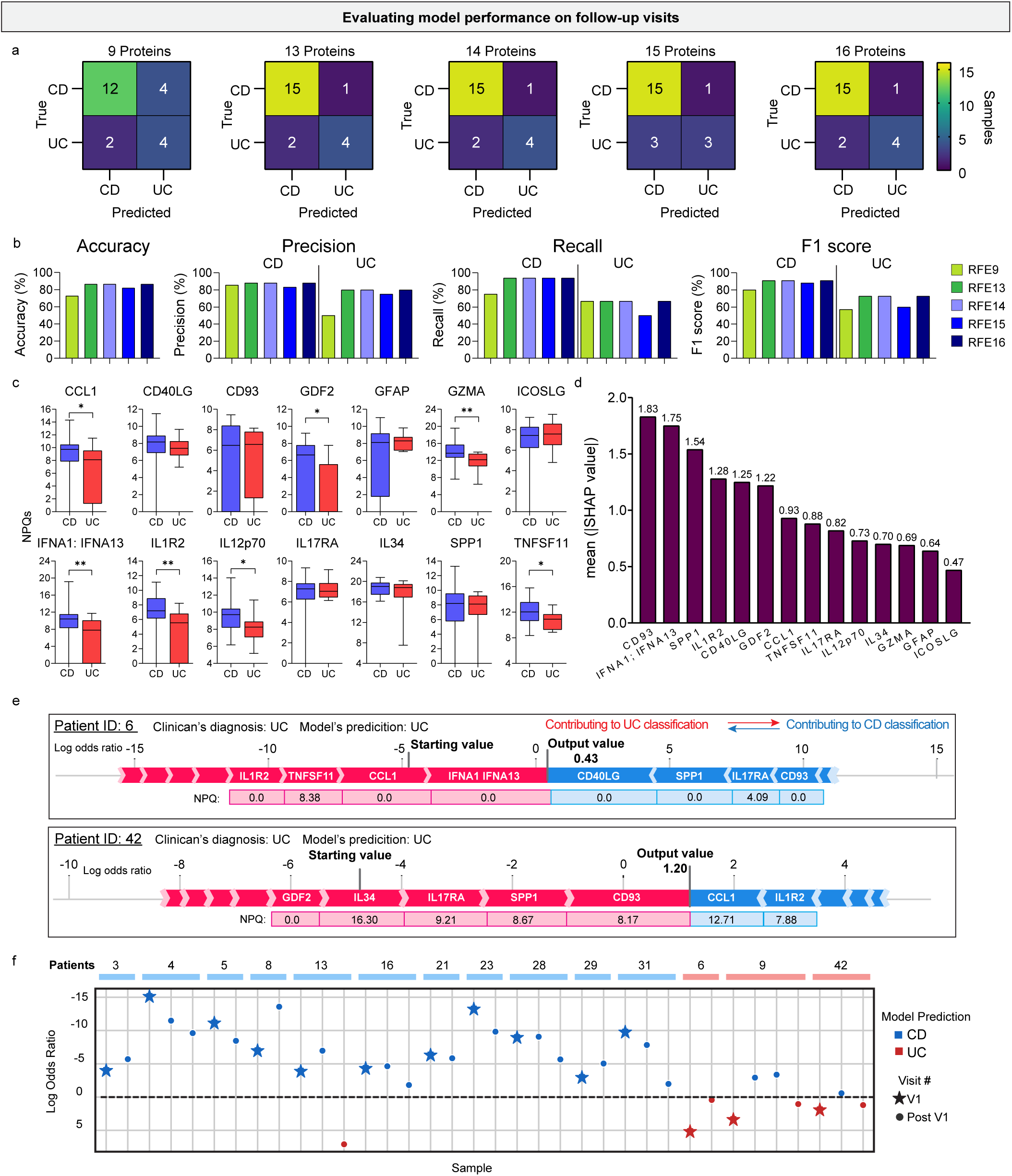
Applying the machine learning model to predict CD and UC diagnosis in samples from patient follow-up visits. a) Confusion matrices showing the agreement between clinician IBD diagnosis (true) and model IBD diagnosis (predicted) for the 5 different RFE models in follow-up visit data. b) Summary bar charts of accuracy, precision, recall, and F1 scores for each model when applied to follow-up visit data. c) Box plots of the 14 proteins used in the RFE model, showing the distribution of normalized protein values (NPQ) in visit 1 CD patient samples (blue, n=40) and UC patient samples (red, n=12). Significance was assessed using a Mann-Whitney test. * p value <0.05, ** p value <0.01. d) Bar graph showing the mean absolute SHAP values across each protein for the follow-up visit samples. e) SHAP force plots showing impact of individual proteins on model’s IBD diagnosis across two samples. Red bars indicate protein is contributing to a UC classification while blue bars indicate protein is contributing to a CD contribution. The size of the bars represents the amount the protein is contributing to the diagnosis. Samples with an output value less than 0 are predicted to be CD, samples with an output value greater than 0 are predicted to be UC. NPQ values for each protein listed are shown below. f) Log odds ratio for patient samples from multiple visits. Stars indicate the value at the initial collection and circles at any follow-up visits. The top patients’ bar indicates which samples were collected from the same patient while the bar color indicates the clinician’s diagnosis (CD: blue, UC: red).

The 14 proteins in the final model are shown with their abundance distributions across CD and UC samples in the training set (Figure 4c). Interestingly, half of these proteins were not statistically different between the two groups when evaluated individually, meaning they would have been overlooked in conventional univariate differential abundance analysis (Figure 2). This underscores the advantage of machine learning approaches, which can identify informative combinations of features that may not be individually significant but together enhance classification power.

To better understand model decision-making, we applied SHapley Additive exPlanations (SHAP) analysis, which highlighted CD93, IFNA1;IFNA13, and SPP1 as the top contributors to model predictions in follow-up visits (Figure 4d). Patient-level force plots further demonstrated how the influence of specific proteins varied between individuals (Figure 4e), reflecting underlying biological heterogeneity within the cohort. For example, in Patient 6 IFNA1;IFNA13 was the top protein supporting a UC classification because of its low protein level, while Patient 42, who was also diagnosed as UC, relied on different proteins entirely to achieve that classification. Such variability reinforces the need for multi-protein panels rather than reliance on a single biomarker.

We also assessed model consistency across repeated samples from the same patient. Of the 14 individuals who contributed multiple visits, only three had a sample misclassified (Figure 4f). Patient 9 had the highest misclassification rate (two of four samples), with correct predictions at Visits 1 and 4 but incorrect results at Visits 2 and 3. Disease activity levels did not explain this pattern, as both correctly and incorrectly classified visits included moderate and quiescent states. Instead, longitudinal shifts in protein abundance appeared more relevant. Specifically, Visits 2 and 3 showed higher CD93 and lower CCL1 and TNFSF11/RANKL compared to other time points (Supplementary Figure 5). This pattern coincided with the initiation of infliximab therapy between the first and second visits, potentially explaining the reduction in TNFSF11/RANKL (RANKL) given its reported association with TNF-α levels.

### Determination of Functional Protein Groups in IBD using Salivary Proteomics

The differentially abundant proteins in our model could be functionally stratified into two predictive groups based on their relative abundance patterns across diagnoses. Group 1, consisting of IL34, TNFSF11/RANKL, IFNA1;IFNA13, IL12p70, GZMA, IL1R2, CCL1, and GDF2, tended to predict Crohn’s disease when present at higher levels and ulcerative colitis when expressed at lower levels. Functional network analysis using STRING revealed biologically meaningful associations within this group, including co-expression links between IL34 and TNFSF11/RANKL as well as between TNFSF11/RANKL and the cytotoxic effector GZMA (Supplementary Figure 6a). These relationships suggest that Group 1 proteins may participate in interconnected inflammatory and immune signaling modules, potentially bridging myeloid–osteoclast axis activity (IL34–RANKL) with cytotoxic granule pathways (RANKL–GZMA), which could contribute to the immunopathological distinctions between pediatric Crohn’s disease and ulcerative colitis.

Group 2, composed of IL17RA, CD40LG, SPP1, CD93, ICOSLG, and GFAP, showed the opposite trend, predicting ulcerative colitis when highly abundant and Crohn’s disease when present at lower levels. This group highlights a distinct set of pathways involving Th17 signaling, costimulatory ligand interactions, extracellular matrix remodeling, and/or vascular activation responses that may differentiate ulcerative colitis–associated inflammatory mechanisms from those observed in Crohn’s disease.

When applying this framework to a longitudinal case (Patient 9) with four visits over one year, the most consistently expressed proteins in Group 1 were IL34, GZMA, and GDF2, while in Group 2, the most stable signals came from ICOSLG, GFAP, and CD40LG, suggesting potential biomarkers for tracking disease phenotype stability or transition over time (Supplementary Figure 5).

### Application of the Model to Indeterminate IBD Cases (IBD-U)

Following the development and testing of the 14-protein model on samples with confirmed CD or UC diagnoses, we next assessed its performance on a set of indeterminate inflammatory bowel disease cases (IBD-U). These cases represent a diagnostically challenging subgroup in which patients exhibit clinical features of IBD but lack sufficient endoscopic, histological, or imaging evidence to classify them definitively as CD or UC. The cohort consisted of eight IBD-U samples from five patients, with a maximum of two visits per patient (Figure 5a).

**Figure 5:**
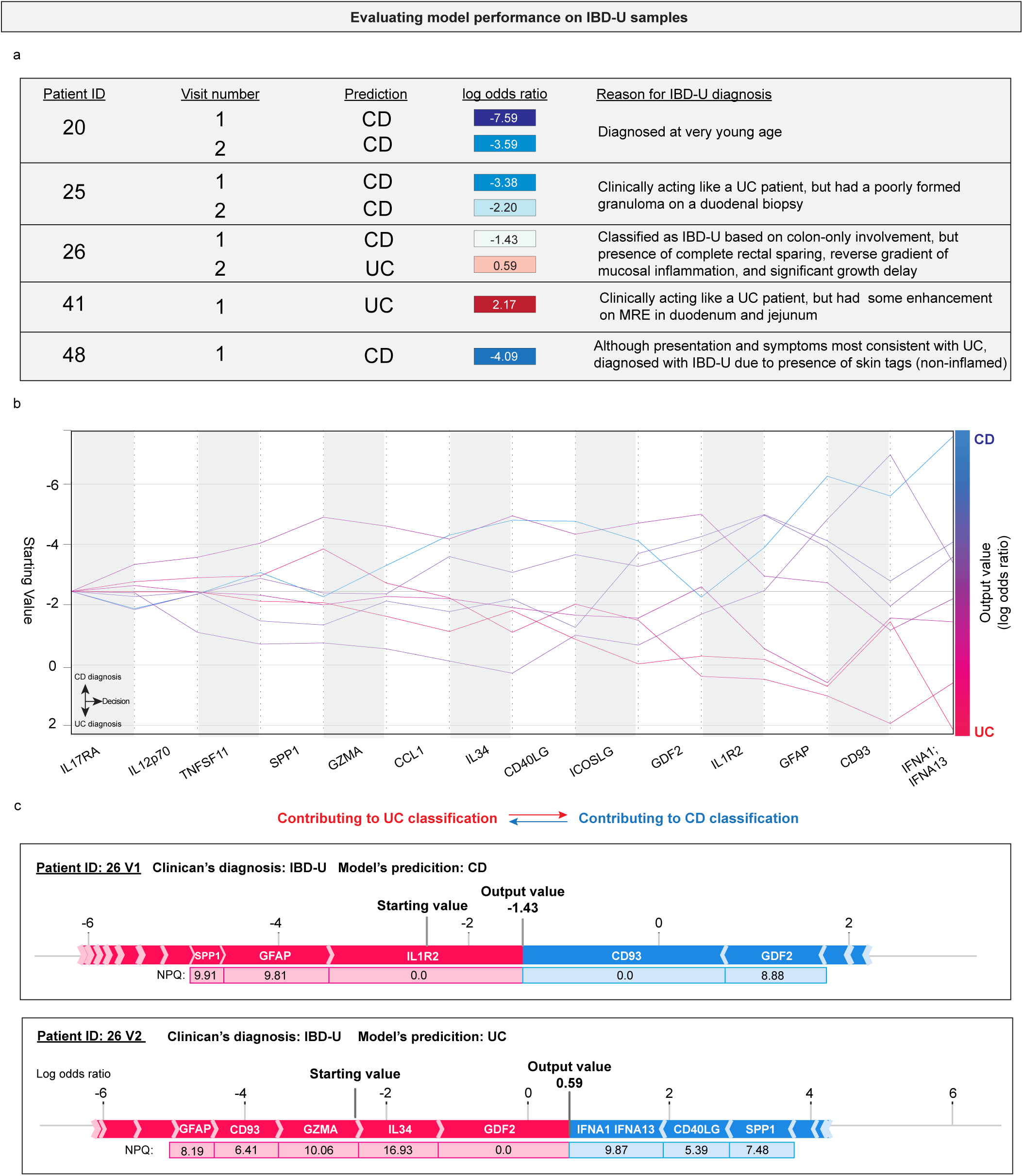
Predicting IBD diagnosis of clinically diagnosed IBD-U patients. a) Chart showing model’s prediction for eight samples from six clinical diagnosed IBD-U patients. Log odds ratio indicates the confidence in the prediction. Negative values indicate final prediction of CD while positive values indicate a final prediction of UC. Reason for IBD-U diagnosis is included for each patient. b) SHAP decision plot of each sample showing impact of each protein on the model’s final prediction. The y-value indicates the log odds ratio, with positive values (red) indicating samples were diagnosed as UC, while negative values (blue) indicate samples were diagnosed as CD. Each sample prediction starts from the same starting value and changes based on the 14 proteins in the RFE model. Larger slopes indicate greater impact of that protein in the final prediction. c) SHAP force plots showing impact of individual proteins on model’s IBD diagnosis across two samples from one patient. Red bars indicate the protein is contributing to a UC classification while blue bars indicate the protein is contributing to a CD classification. The size of the bars represents the amount the protein is contributing to the diagnosis. Samples with an output value less than 0 are diagnosed as CD, samples with an output value greater than 0 are diagnosed as UC. NPQ values are shown below the indicated protein.

For each sample, we recorded patient identifiers, reason for IBD-U diagnosis, the model’s predicted classification, and the corresponding log odds ratio which indicate the model’s confidence. In the context of our model, a negative log odds ratio indicates a prediction of CD, a positive value indicates a prediction of UC, and a higher absolute log odds ratio reflects greater certainty in the model’s classification, while values near zero suggest ambiguity. The majority of multi-visit patients (two out of three) received consistent model predictions across their time points, suggesting some stability in the proteomic signals for those individuals. However, one patient (Patient 26) displayed conflicting predictions between visits, with low absolute log odds ratios indicating that the model lacked strong confidence in either classification. Such low-confidence predictions highlight the potential for underlying biological or disease state variability within an individual that may not be fully captured by a single time point’s proteomic profile.

To better understand how the model reached its predictions in IBD-U cases, we examined SHapley Additive exPlanation (SHAP) decision plots for all eight samples (Figure 5b). These plots display the cumulative contribution of each protein, ordered from overall least to most influential in the model’s decision process. Here we can see if the model’s prediction remained consistent with the addition of another protein or if one or more proteins caused an intense shift in overall prediction. These abrupt directional changes in the decision pathway indicate that specific proteins can exert strong individual influence, occasionally overriding the broader multi-protein signal.

We further investigated conflicting cases (Patient 26) using force plots for selected samples (Figure 5c), which provide a more granular visualization of how each protein’s NPQ value contributes to the final prediction. In both visits for Patient 26, certain proteins acted as strong CD drivers despite the majority of markers aligning with a UC diagnosis. CD93 was the top protein contributing to a CD diagnosis in visit 1 but shifted to predict UC in visit 2 due to an increase its salivary protein level. This conflicting signal pattern and varying protein amount could be reflective of concurrent inflammatory processes, atypical immune activation, or treatment-related effects occurring at the time of sample collection. Such findings underscore the importance of integrating multiple layers of information, such as SHAP contribution patterns, log odds ratios, and longitudinal trends, when interpreting model outputs for diagnostically ambiguous patients. Ultimately, these insights suggest that the model, when coupled with interpretable machine learning techniques, has potential not only for classifying established IBD cases but also for providing clinically informative guidance in complex, indeterminate scenarios where traditional diagnostic pathways are inconclusive.

## Discussion

Inflammatory bowel diseases (IBD) can present with a wide range of debilitating symptoms affecting multiple aspects of a patient’s daily life. As such, the primary goal of clinicians is to rapidly assess and treat affected individuals to alleviate discomfort, prevent disease progression, and ultimately improve quality of life^31^. However, the clinical management of IBD is complicated by the substantial complexity and heterogeneity of these diseases, which is driven by a multifaceted interplay between genetic susceptibility, immune dysregulation, environmental exposures, and the gut microbiome^32^. These factors contribute to overlapping clinical features between Crohn’s disease (CD) and ulcerative colitis (UC). Nevertheless, accurate diagnosis, management, and monitoring of IBD cases is crucial, as therapeutic approaches and prognostic outcomes differ. This diagnostic challenge is further amplified in pediatric patients, who often exhibit more extensive intestinal involvement, greater disease severity, and faster progression compared to adults^33^.

Against this backdrop, there is a pressing need for novel, minimally invasive diagnostic tools that can accurately differentiate IBD subtypes in children, reduce diagnostic uncertainty in IBD-U, and provide actionable insights that complement conventional endoscopic, histologic, and imaging-based evaluations. While there are limited studies utilizing saliva as a liquid biopsy for IBD, especially in pediatric cases, surveys of adult patients and parents of pediatric patients found saliva-based test to be the preferred choice for disease monitoring when given a list of current options (blood, fecal, colonoscopy, etc.) and potential future options (saliva, urine, breath)^34,35^. We found saliva to be a suitable biofluid for this type of study as it was viewed as non-invasive with easy collection, leading to high patient recruitment, and required only 30ul to be assayed on the NULISAseq platform. Additionally, due to its easy collection and processing compared to blood and stool, saliva may be extremely beneficial for future large-scale studies in which longitudinal biosample collection is required.

Several proteins in our 14-marker panel have been implicated in IBD and inflammatory pathways, including GDF2/BMP9, IL34, and CD40LG^36-38^, though few have been used as CD- or UC-specific biomarkers^39^. Within our model, these proteins can be stratified into two functional groups with distinct predictive trends. Group 1 (IL34, TNFSF11/RANKL, IFNA1;IFNA13, IL12p70, GZMA, IL1R2, CCL1, GDF2) predicted Crohn’s disease when present at higher levels and ulcerative colitis when expressed at lower levels, while Group 2 (IL17RA, CD40LG, SPP1, CD93, ICOSLG, GFAP) predicted ulcerative colitis when abundant and Crohn’s disease when reduced. KEGG pathway analysis revealed that many Group 1 proteins (IL34, TNFSF11/RANKL, IFNA1, IL12p70, IL1R2, CCL1, GDF2) participate in cytokine–cytokine receptor interactions (padj = 5.1 × 10^-7), while some Group 2 proteins (CD40LG, ICOSLG) are associated with the intestinal immune network for IgA production (padj=1.3×10^-2).

Considering the oral cavity as the focus of this project for sampling, the biological relevance of these functional group findings is underscored by the functional roles of certain Group 1 members, particularly IL34 and TNFSF11/RANKL, which are potent inducers of osteoclastogenesis^40,41^. While these cytokines are known to be elevated within oral fluids during periodontal diseases^42^, the young age of our pediatric cohort and the fact that periodontitis is often a disease of younger to middle-aged adult makes localized periodontal pathology an unlikely explanation. Instead, their presence likely reflects systemic inflammatory activity in IBD^11^. Oral lesions, especially gingival inflammation and periodontitis, have been increasingly recognized extraintestinal manifestations of IBD^43^, and the salivary detection of osteoclastogenic cytokines may signal a broader axis connecting systemic inflammation, oral tissue pathology, and skeletal health in younger patients that may become clearer in those with active periodontal disease in future studies.

The computational strategy for this work was selected as it has demonstrated the ability to optimize both accuracy and interpretability^44^. Recursive feature elimination (RFE) was applied to iteratively remove less informative proteins, yielding a compact, highly predictive panel. From a clinical perspective, a reduced biomarker set lowers assay cost and turnaround time, supporting potential translation into practice. RFE also unexpectedly streamlined quality control. Standard post-normalization QC guidelines suggests removing proteins below the limit of detection (LOD) in more than 50% of samples; however, this approach excluded two proteins (GDF2 and IL17RA) that proved important for classification. For example, IL17RA was below the LOD in 66.6% of UC samples but only 47.5% of CD samples, suggesting that detection frequency itself may be informative for subtype discrimination. Thus, proteins below LOD should not automatically be excluded without careful review.

Given the costs associated with proteomic profiling, our initial sample size was modest. We therefore used leave-one-out cross-validation, which maximizes available training data while still providing robust accuracy estimates. Expanding the cohort in future work will allow refinement of the model and incorporation of additional clinical parameters such as disease activity and treatment status, which may influence proteomic profiles. The goal is to define the smallest possible set of proteins that delivers high predictive accuracy, enabling development of a rapid, affordable clinical test. Larger datasets with serial samples could also enable longitudinal analyses, including detection of shifts in proteomic patterns that precede clinical relapse or treatment failure.

A common limitation of machine learning models is their “black box” nature, which can hinder clinical adoption. To address this, we incorporated SHapley Additive exPlanations (SHAP) to identify the contribution of each protein to individual predictions. Force and decision plots illustrated how each marker shifted model output from a baseline log-odds score toward the final classification. Although some proteins were generally more influential than others, their impact varied across patients, sometimes even switching direction depending on protein abundance. This variability reflects biological heterogeneity and reinforces the utility of multi-marker approaches. Our analysis also revealed that the model baseline was slightly biased toward predicting CD, which may have contributed to the lower UC recall and F1 scores in follow-up samples. Increasing UC representation in the training set should help balance this tendency.

The inflammatory panel used in this study was selected for relevance to IBD but is not disease-specific, meaning elevated levels could also arise from unrelated inflammatory conditions. The SHAP analysis, which sometimes revealed conflicting protein-level predictions, underscores the importance of comprehensive clinical evaluation alongside biomarker testing. In our cohort, no patients received vaccines within two weeks of sampling, and none had IBD-related surgery within one month. While collectors were instructed to check for overt oral inflammation before sampling, this was not consistently documented, potentially due to limited familiarity with oral examination. Future protocols will include targeted training for clinical staff to ensure standardized oral health assessments.

Our model achieved 86% accuracy in predicting IBD subtype in annotated cases not used for training. We anticipate similar performance in IBD-U patients, though longitudinal follow-up will be needed for confirmation. In clinical practice, each model output would be accompanied by SHAP-based interpretability plots and a log-odds confidence score, enabling clinicians to integrate biomarker results with patient history, endoscopic findings, and other diagnostic data to guide individualized management decisions. Ultimately, the integration of salivary proteomics with explainable machine learning offers a promising path toward minimally invasive, accurate, and interpretable diagnostics for pediatric IBD. This approach could facilitate earlier, more precise classification of disease subtype, reduce reliance on invasive procedures, and support personalized treatment strategies, thereby addressing a critical need for pediatric patients with these debilitating disorders.

## Acknowledgements

We want to thank the team at UNC’s Marsico Lung Institute Respiratory TRACTS Core, especially Mandy Bush and Griselda Portillo, for isolating saliva supernatant, storing and shipping samples. We want to acknowledge the Alamar Biosciences’ Technology Access Program for running the healthy adult pilot study and pediatric IBD samples on the NULISAseq platform. Figures 1a, 1b, 3a, and S4a were made using Biorender.com.

## Ethical Considerations

The protocol (UNC IRB # 23-2319) was reviewed and approved by the institutional review board at the University of North Carolina. The study was conducted in compliance with the Declaration of Helsinki. All participants or their legal guardians provided written, informed consent to participate.

## Author Contributions

For this study, KMB and ASG conceptualized the project. JL and JR designed the algorithm. JR implemented the algorithm and conducted benchmarking experiments. TW and BTR performed manual assessment of clinical outcome. ASG, AG, and KC supported the recruitment of patients and collected data. BTR, JR, KMB, and JL performed experimental and/or bioinformatic analyses that supported project development. JR, JL, BTR, KMB, and ASG wrote the original draft; BTR, JR, AG, TW, KC, JL, ASG, and KMB critically reviewed and edited the final manuscript.

## Data Availability

Data and code can be found at: https://github.com/VCU-Bioinformatics-Core/20241011.kevin_byrd.ibd_nulisa

## Supplementary Material Legends

**Supplementary Table 1:** | List of 250 proteins included in the NULISAseq Inflammation Panel.

**Supplemental Figure 1: Pilot data examining sample volume needed to reach proteins’ limit of detection (LOD)**. | Heatmap of protein level (NPQ) - limit of detection (LOD) for 10ul and 30ul saliva samples collected at the same time from the same healthy adult. Black bars indicate proteins that did not reach the LOD in that sample. The number of proteins at or above the LOD is listed below.

**Supplemental Figure 2: Salivary calprotectin levels in active and inactive IBD patients**. | Box plot of S100A9 protein level (NULISA protein quantification units (NPQ) in IBD patients with active or inactive disease.

**Supplemental Figure 3: Additional box plots of statistically significant differential abundant proteins not included in Figure 2c. |** a) Box plots of statistically significant (p-value <0.05) differential abundant proteins when CD is compared to UC. Box plots show the distribution of protein levels (NULISA protein quantification units (NPQ)) in all CD patient samples (blue) and UC patient samples (red). Significance was determined using differential abundance analysis. * p value <0.05, ** p value <0.01.

**Supplemental Figure 4: Reanalysis using Limit of Detection (LOD) cutoff.** | a) Overview of LOD cutoff reanalysis. b) Principal component analysis using the 230 proteins that passed the LOD cutoff across all Crohn’s (CD) and ulcerative colitis (UC) samples, colored by IBD diagnosis. c) Heatmap with hierarchical clustering of samples showing 230 standardized protein levels from the NULISAseq Inflammation Panel. Corresponding IBD diagnosis and disease activity for each sample is shown. d) Confusion matrices showing the agreement between clinician IBD diagnosis (true) and model IBD diagnosis (predicted) in first visit data for 5 different RFE models generated using the 230 proteins above LOD cutoff, with accuracy and performance statistics listed for each model. e) Summary bar charts of accuracy, precision, recall, and F1 scores for each model when applied to visit 1 data. f) Confusion matrices showing the agreement between clinician IBD diagnosis (true) and model IBD diagnosis (predicted) in follow-up data for 5 different 230 protein RFE models generated using first visit data, with accuracy and performance statistics listed for each model. g) Summary bar charts of accuracy, precision, recall, and F1 scores for each model when applied to post visit 1 data. Illustrations from a) were created with BioRender.com

**Supplemental Figure 5: Heatmap of model’s protein levels across all samples from patient 9.** | NULISA protein quantification (NPQ) levels for all 14 proteins used in the RFE model, across samples collected at each of patient 9’s visit.

**Supplemental Figure 6: Results from STRING analysis on proteins in machine learning model.** | a) STRING generated network summary proteins from Group 1 proteins (IL34, TNFSF11/RANKL, IFNA1;IFNA13, IL12p70, GZMA, IL1R2, CCL1, GDF2. b) STRING generated network summary proteins from Group 2 proteins (IL17RA, CD40LG, SPP1, CD93, ICOSLG, GFAP).

## REFERENCES

1 Roda, G. et al. Crohn’s disease. Nat Rev Dis Primers 6, 22 (2020). 10.1038/s41572-020-0156-2

2 Kobayashi, T. et al. Ulcerative colitis. Nat Rev Dis Primers 6, 74 (2020). 10.1038/s41572-020-0205-x

3 Bishop, J., Lemberg, D. A. & Day, A. Managing inflammatory bowel disease in adolescent patients. Adolesc Health Med Ther 5, 1–13 (2014). 10.2147/ahmt.S37956

4 Moeeni, V. & Day, A. S. Impact of Inflammatory Bowel Disease upon Growth in Children and Adolescents. ISRN Pediatr 2011, 365712 (2011). 10.5402/2011/365712

5 Guariso, G. & Gasparetto, M. Treating children with inflammatory bowel disease: Current and new perspectives. World J Gastroenterol 23, 5469–5485 (2017). 10.3748/wjg.v23.i30.5469

6 Gaitsch, H., Franklin, R. J. M. & Reich, D. S. Cell-free DNA-based liquid biopsies in neurology. Brain 146, 1758–1774 (2023). 10.1093/brain/awac438

7 Ma, L. et al. Liquid biopsy in cancer current: status, challenges and future prospects. Signal Transduct Target Ther 9, 336 (2024). 10.1038/s41392-024-02021-w

8 Song, M., Bai, H., Zhang, P., Zhou, X. & Ying, B. Promising applications of human-derived saliva biomarker testing in clinical diagnostics. Int J Oral Sci 15, 2 (2023). 10.1038/s41368-022-00209-w

9 Humphrey, S. P. & Williamson, R. T. A review of saliva: normal composition, flow, and function. J Prosthet Dent 85, 162–169 (2001). 10.1067/mpr.2001.113778

10 Shazib, M. A., Byrd, K. M. & Gulati, A. S. Diagnosis and Management of Oral Extraintestinal Manifestations of Pediatric Inflammatory Bowel Disease. J Pediatr Gastroenterol Nutr 74, 7–12 (2022). 10.1097/mpg.0000000000003302

11 Byrd, K. M. & Gulati, A. S. The "Gum-Gut" Axis in Inflammatory Bowel Diseases: A Hypothesis-Driven Review of Associations and Advances. Front Immunol 12, 620124 (2021). 10.3389/fimmu.2021.620124

12 Deng, Y. T. et al. Atlas of the plasma proteome in health and disease in 53,026 adults. Cell 188, 253–271.e257 (2025). 10.1016/j.cell.2024.10.045

13 Oh, H. S. et al. A cerebrospinal fluid synaptic protein biomarker for prediction of cognitive resilience versus decline in Alzheimer’s disease. Nature medicine 31, 1592–1603 (2025). 10.1038/s41591-025-03565-2

14 Feng, W. et al. NULISA: a proteomic liquid biopsy platform with attomolar sensitivity and high multiplexing. Nat Commun 14, 7238 (2023). 10.1038/s41467-023-42834-x

15 Topol, E. J. The revolution in high-throughput proteomics and AI. Science (New York, N.Y.) 385, eads5749 (2024). 10.1126/science.ads5749

16 Kalla, R. et al. Serum proteomic profiling at diagnosis predicts clinical course, and need for intensification of treatment in inflammatory bowel disease. J Crohns Colitis 15, 699–708 (2021). 10.1093/ecco-jcc/jjaa230

17 Grännö, O. et al. Preclinical Protein Signatures of Crohn’s Disease and Ulcerative Colitis: A Nested Case-Control Study Within Large Population-Based Cohorts. Gastroenterology 168, 741–753 (2025). 10.1053/j.gastro.2024.11.006

18 Gorelik, M. G. et al. Improving Differentiation of Crohn’s Disease and Ulcerative Colitis Proteomes through Protein-Wide Association Study Feature Selection in Machine Learning. medRxiv (2024). 10.1101/2024.11.13.24316854

19 Salomon, B. et al. Characterization of Inflammatory Bowel Disease Heterogeneity Using Serum Proteomics: A Multicenter Study. J Crohns Colitis 19 (2025). 10.1093/ecco-jcc/jjae169

20 Vuijk, S. A., Camman, A. E. & de Ridder, L. Considerations in Paediatric and Adolescent Inflammatory Bowel Disease. J Crohns Colitis 18, ii31-ii45 (2024). 10.1093/ecco-jcc/jjae087

21 Kelsen, J. & Baldassano, R. N. Inflammatory bowel disease: the difference between children and adults. Inflamm Bowel Dis 14 Suppl 2, S9–11 (2008). 10.1002/ibd.20560

22 Virtanen, P. et al. SciPy 1.0: fundamental algorithms for scientific computing in Python. Nat Methods 17, 261–272 (2020). 10.1038/s41592-019-0686-2

23 Gu, Z. Complex heatmap visualization. Imeta 1, e43 (2022). 10.1002/imt2.43

24 Ritchie, M. E. et al. limma powers differential expression analyses for RNA-sequencing and microarray studies. Nucleic Acids Res 43, e47 (2015). 10.1093/nar/gkv007

25 LemaÃŽtre, G., Nogueira, F. & Aridas, C. K. Imbalanced-learn: A python toolbox to tackle the curse of imbalanced datasets in machine learning. Journal of machine learning research 18, 1–5 (2017).

26 Pedregosa, F. et al. Scikit-learn: Machine learning in Python. the Journal of machine Learning research 12, 2825–2830 (2011).

27 Lundberg, S. M. & Lee, S.-I. A unified approach to interpreting model predictions. Advances in neural information processing systems 30 (2017).

28 Bos, V. et al. Salivary Calprotectin Is not a Useful Biomarker to Monitor Disease Activity in Patients with Inflammatory Bowel Disease. J Gastrointestin Liver Dis 31, 283–289 (2022). 10.15403/jgld-4215

29 Caron, B. et al. Gastroenterological safety of IL-17 inhibitors: a systematic literature review. Expert Opin Drug Saf 21, 223–239 (2022). 10.1080/14740338.2021.1960981

30 Ward, D. et al. Tumor Necrosis Factor Inhibitors in Inflammatory Bowel Disease and Risk of Immune Mediated Inflammatory Diseases. Clin Gastroenterol Hepatol 22, 135–143.e138 (2024). 10.1016/j.cgh.2023.06.025

31 Ye, B. D. & Travis, S. Improving the quality of care for inflammatory bowel disease. Intest Res 17, 45–53 (2019). 10.5217/ir.2018.00113

32 Chang, J. T. Pathophysiology of Inflammatory Bowel Diseases. N Engl J Med 383, 2652–2664 (2020). 10.1056/NEJMra2002697

33 Kim, J. & Ye, B. D. Successful transition from pediatric to adult care in inflammatory bowel disease: what is the key? Intest Res 17, 24–35 (2019). 10.5217/ir.2018.00128

34 Ho, S. S. C., Keenan, J. I. & Day, A. S. Patient perceptions of current and potential inflammatory bowel disease diagnostic and monitoring tests. Intern Med J 52, 1196–1202 (2022). 10.1111/imj.15298

35 Ho, S. S. C., Keenan, J. I. & Day, A. S. Parent Perspectives of Diagnostic and Monitoring Tests Undertaken by Their Child with Inflammatory Bowel Disease. Pediatr Gastroenterol Hepatol Nutr 24, 19–29 (2021). 10.5223/pghn.2021.24.1.19

36 Xie Z. et al. Recent developments on BMPs and their antagonists in inflammatory bowel diseases. Cell Death Discov. 9, 210 (2023).10.1038/s41420-023-01520-z

37 Monteleone, G., Franzè, E., Troncone, E., Maresca, C. & Marafini, I. Interleukin-34 Mediates Cross-Talk Between Stromal Cells and Immune Cells in the Gut. Front Immunol 13, 873332 (2022). 10.3389/fimmu.2022.873332

38 Senhaji, N., Kojok, K., Darif, Y., Fadainia, C. & Zaid, Y. The Contribution of CD40/CD40L Axis in Inflammatory Bowel Disease: An Update. Front Immunol 6, 529 (2015). 10.3389/fimmu.2015.00529

39 Zhou, P. et al. Unveiling the hidden dance: SPP1[+[macrophages identified in ulcerative colitis reveal crosstalk with CHI3L1[+[fibroblasts. J Transl Med 23, 567 (2025). 10.1186/s12967-025-06565-5

40 Chen, Z., Buki, K., Vääräniemi, J., Gu, G. & Väänänen, H. K. The critical role of IL-34 in osteoclastogenesis. PLoS One 6, e18689 (2011). 10.1371/journal.pone.0018689

41 Park, J. H., Lee, N. K. & Lee, S. Y. Current Understanding of RANK Signaling in Osteoclast Differentiation and Maturation. Mol Cells 40, 706–713 (2017). 10.14348/molcells.2017.0225

42 Bozkurt Doğan, Ş., Öngöz Dede, F., Ballı, U. & Sertoğlu, E. Emerging roles of Interleukin-34 together with receptor activator of nuclear factor-kB ligand and osteoprotegerin levels in periodontal disease. Cytokine 144, 155584 (2021). 10.1016/j.cyto.2021.155584

43 Ribaldone, D. G. et al. Oral Manifestations of Inflammatory Bowel Disease and the Role of Non-Invasive Surrogate Markers of Disease Activity. Medicines (Basel) 7 (2020). 10.3390/medicines7060033

44 Yee, J. Y. et al. Predicting antipsychotic responsiveness using a machine learning classifier trained on plasma levels of inflammatory markers in schizophrenia. Transl Psychiatry 15, 51 (2025). 10.1038/s41398-025-03264-z

