## Supplementary Table 1 for "Interpretable machine learning applied to high-dimensional salivary proteomics accurately classifies pediatric inflammatory bowel diseases"

| NULISAseq Inflammation Panel 250 | | | | | | | | | | |
| --- | --- | --- | --- | --- | --- | --- | --- | --- | --- | --- |
| AGER | CCL23 | CD83 | CXCL13 | GFAP | IL-13 | IL-22 | IL-9 | MMP9 | TAFA5 | TNFSF10 |
| AGRP | CCL24 | CD93 | CXCL14 | GRN | IL-13RA2 | IL-23A\|EBI3 | IRAK4 | MPO | TEK | TNFSF11 |
| ANGPT1 | CCL25 | CEACAM5 | CXCL16 | GZMA | IL-15 | IL-23A\|IL-12B | KDR | MUC16 | TGFB1 | TNFSF12 |
| ANGPT2 | CCL26 | CHI3L1 | CXCL2 | GZMB | IL-15RA | IL-24 | KITLG | NAMPT | TGFB3 | TNFSF13 |
| ANXA1 | CCL27 | CLEC4A | CXCL3 | HAVCR1 | IL-16 | IL-27\|EBI3 | KLRK1 | NCR1 | THBS2 | TNFSF13B |
| AREG | CCL28 | CNTF | CXCL5 | HGF | IL-17A | IL-2RA | KNG1 | NGF | THPO | TNFSF14 |
| BDNF | CCL3 | CRP | CXCL6 | HLA-DRA | IL-17A\|IL-17F | IL-2RB | LAG3 | NTF3 | TIMP1 | TNFSF15 |
| BMP7 | CCL4 | CSF1 | CXCL8 | ICAM1 | IL-17B | IL-32 | LAMP3 | OSM | TIMP2 | TNFSF18 |
| BST2 | CCL5 | CSF1R | CXCL9 | ICOSLG | IL-17C | IL-33 | LCN2 | PDCD1 | TLR3 | TNFSF4 |
| C1QA | CCL7 | CSF2 | EGF | IFNA1; IFNA13 | IL-17F | IL-34 | LGALS9 | PDCD1LG2 | TNF | TNFSF8 |
| CALCA | CCL8 | CSF2RB | EPO | IFNA2 | IL-17RA | IL-36A | LIF | PDGFA | TNFRSF11A | TNFSF9 |
| CCL1 | CD200 | CSF3 | FASLG | IFNB1 | IL-17RB | IL-36G | LIL-RB2 | PDGFB | TNFRSF11B | TREM1 |
| CCL11 | CD200R1 | CSF3R | FGF19 | IFNG | IL-18 | IL-37 | LTA | PGF | TNFRSF13B | TREM2 |
| CCL13 | CD27 | CST7 | FGF2 | IFNL1 | IL-18BP | IL-3RA | LTA\|LTB | PTX3 | TNFRSF13C | VCAM1 |
| CCL14 | CD274 | CTF1 | FGF21 | IFNL2;IFNL3 | IL-18R1 | IL-4 | MDK | S100A12 | TNFRSF14 | VEGFA |
| CCL15 | CD276 | CTLA4 | FGF23 | IFNW1 | IL-19 | IL-4R | MERTK | S100A9 | TNFRSF17 | VEGFC |
| CCL16 | CD3E | CTSS | FLT1 | IKBKG | IL-1B | IL-5 | MICA | SCG2 | TNFRSF18 | VEGFD |
| CCL17 | CD4 | CX3CL1 | FLT3LG | IL-10 | IL-1R1 | IL-5RA | MICB | SDC1 | TNFRSF1A | VSNL1 |
| CCL19 | CD40 | CXADR | FLT4 | IL-10RB | IL-1R2 | IL-6 | MIF | SELE | TNFRSF1B | VSTM1 |
| CCL2 | CD40LG | CXCL1 | FTH1 | IL-11 | IL-1RL1 | IL-6R | MMP1 | SELP | TNFRSF21 | WNT16 |
| CCL20 | CD46 | CXCL10 | FURIN | IL-12A\|IL-12B | IL-1RN | IL-6ST | MMP12 | SIRPA | TNFRSF4 | WNT7A |
| CCL21 | CD70 | CXCL11 | GDF15 | IL-12B | IL-2 | IL-7 | MMP3 | SLAMF1 | TNFRSF8 |  |
| CCL22 | CD80 | CXCL12 | GDF2 | IL-12RB1 | IL-20 | IL-7R | MMP8 | SPP1 | TNFRSF9 |  |
