## Supplementary figures and images for "Interpretable machine learning applied to high-dimensional salivary proteomics accurately classifies pediatric inflammatory bowel diseases"

### Supplemental Figure 1

Supplementary Figure 1

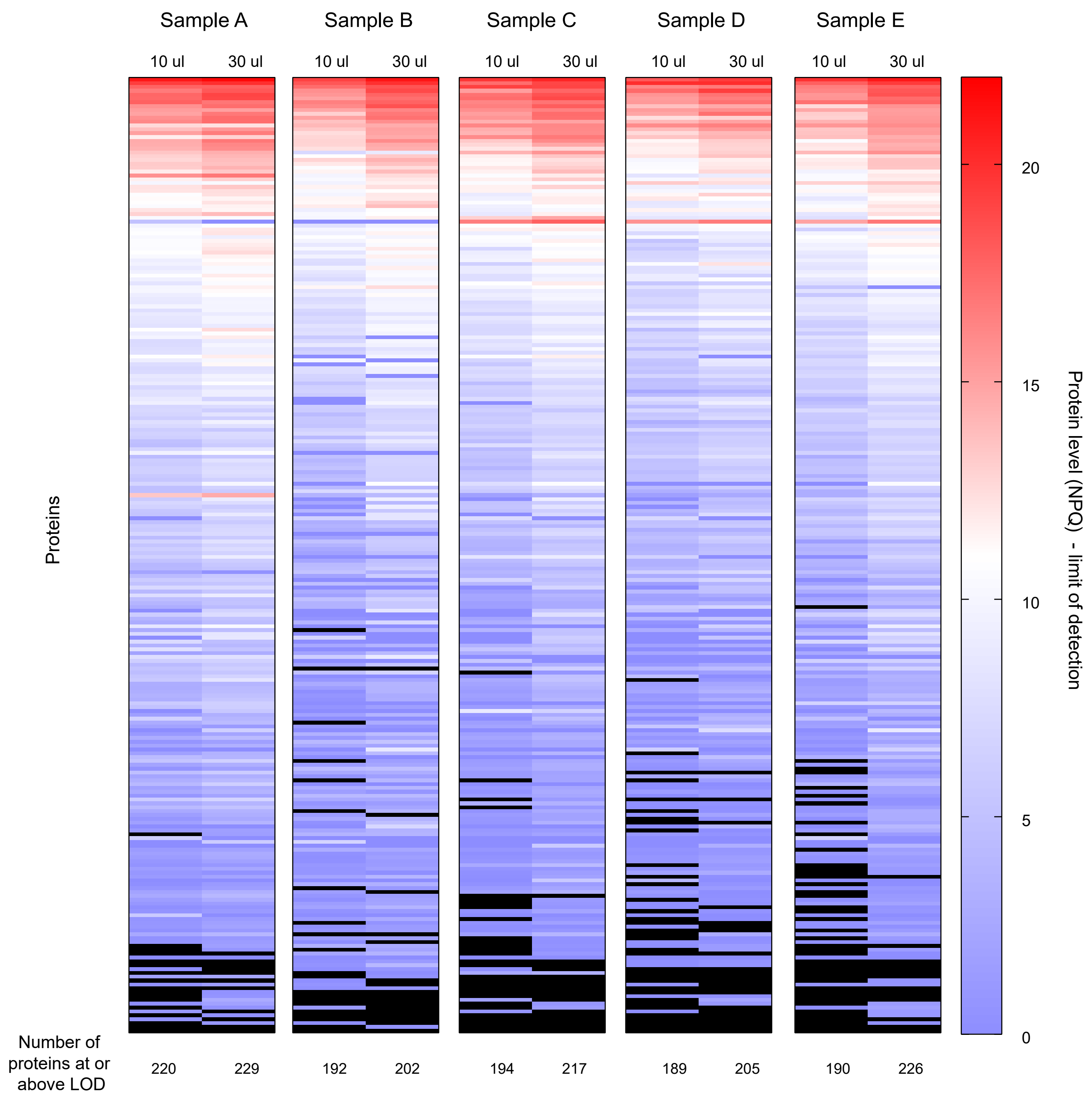

### Supplemental Figure 2

Supplementary Figure 2

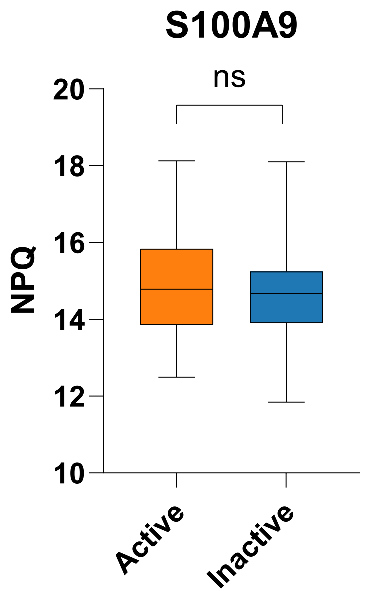

### Supplemental Figure 3

Supplementary Figure 3

**Additional 42 Differentially Abundant Proteins**

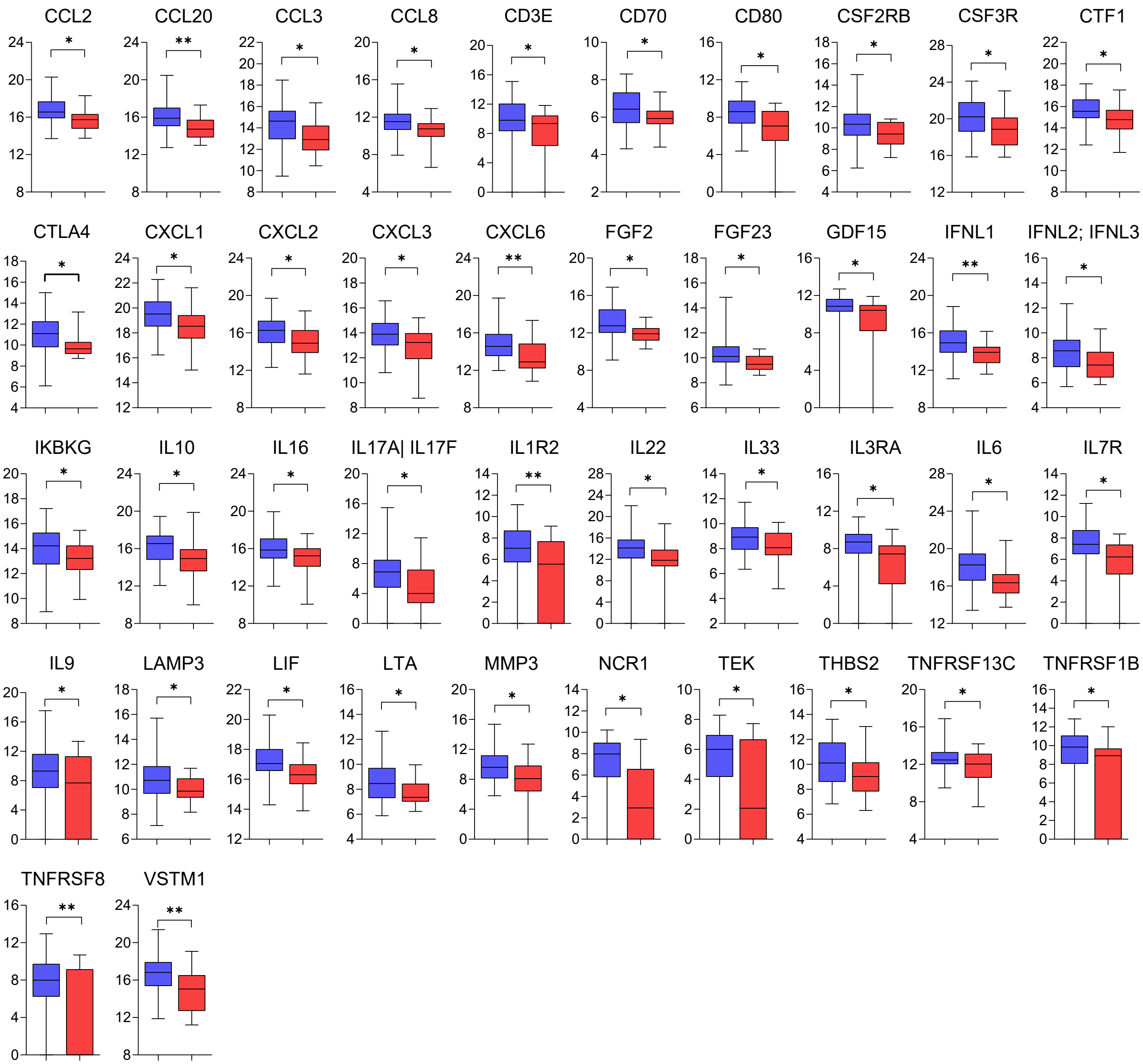

### Supplemental Figure 4

Supplmentary Figure 4

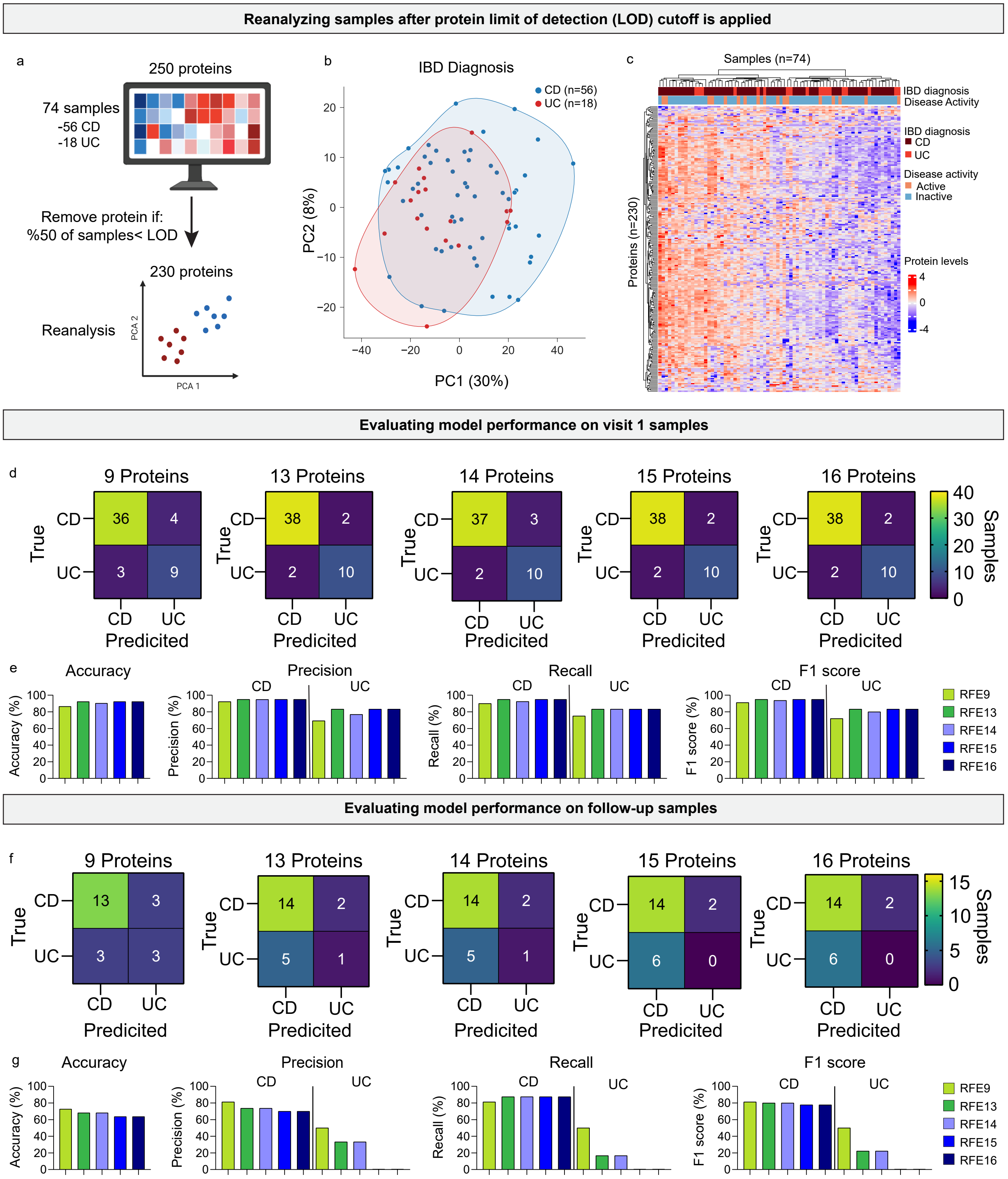

### Supplemental Figure 5

Supplementary Figure 5

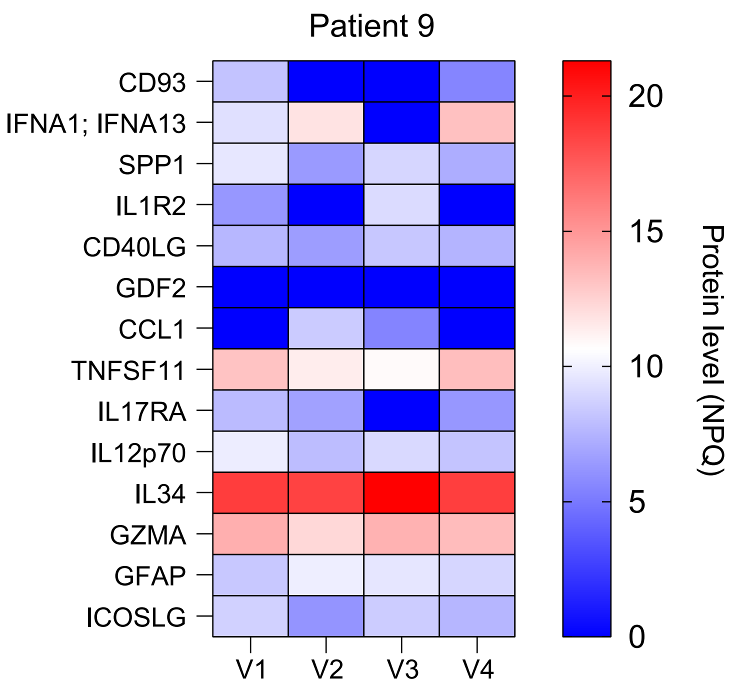

### Supplemental Figure 6

Supplementary Figure 6

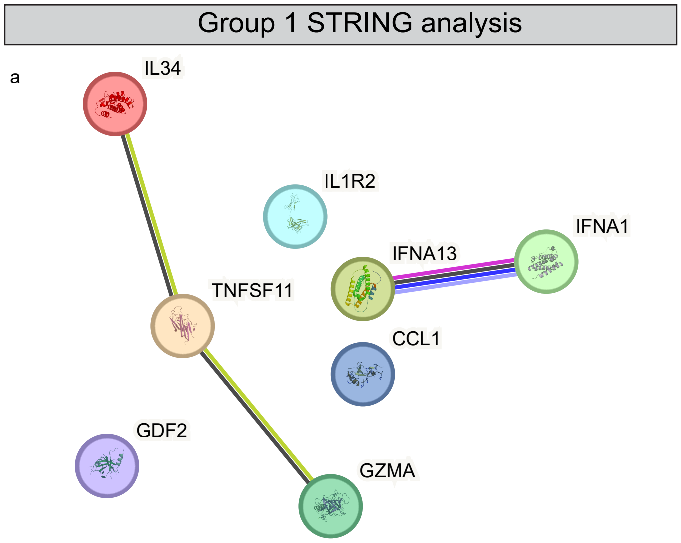


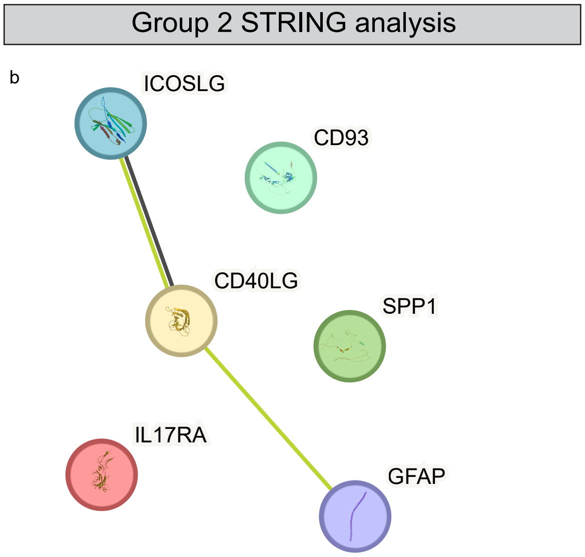
